# Biomarker-guided neuromodulation aids memory in traumatic brain injury

**DOI:** 10.1101/2021.05.18.21256980

**Authors:** Michael J. Kahana, Paul A. Wanda, Youssef Ezzyat, Ethan A. Solomon, Richard Adamovich-Zeitlin, Bradley Lega, Barbara C. Jobst, Robert E. Gross, Kan Ding, Ramon R. Diaz-Arrastia

**Affiliations:** Department of Psychology, University of Pennsylvania; Department of Psychology, Wesleyan University; Department of Neurosurgery, University of Texas Southwestern; Department of Neurology, Dartmouth-Hitchcock Medical Center; Department of Neurosurgery, Emory University; Department of Neurology and Neurotherapeutics, University of Texas Southwestern; Department of Neurology, University of Pennsylvania

## Abstract

Traumatic brain injury (TBI) is a leading cause of cognitive disability in adults, often characterized by marked deficits in episodic memory and executive function. Prior studies have found that direct electrical stimulation of the temporal cortex yielded improved memory in epilepsy patients, but it is not clear if these results generalize to patients with a specific history of TBI. Here we asked whether applying closed-loop, direct electrical stimulation to lateral temporal cortex could reliably improve memory in a TBI cohort. Among a larger group of patients undergoing neurosurgical evaluation for refractory epilepsy, we recruited a subset patients with a history of moderate-to-severe TBI. By analyzing neural data from indwelling electrodes as patients studied and recalled lists of words, we trained personalized machine-learning classifiers to predict momentary fluctuations in mnemonic function in each patient. We subsequently used these classifiers to trigger high-frequency stimulation of the lateral temporal cortex (LTC) at moments when memory was predicted to fail. This strategy yielded a 19% boost in recall performance on stimulated as compared with non-stimulated lists (*P* = 0.012). These results provide a proof-of-concept for using closed-loop stimulation of the brain in treatment of TBI-related memory impairment.

## Introduction

Memory loss resulting from traumatic, infectious, or inflammatory insults to the brain constitutes one of the major health challenges affecting populations worldwide. Disability resulting from traumatic brain injury (TBI), in particular, affects 1-2% of the population and often results in a profound and specific impairment in episodic memory, preventing affected individuals from maintaining a reasonable quality of life. Prior TBI also increases risk for several chronic neurologic complications, including epilepsy *(1)* and neurodegenerative diseases *(2–4)*. The variability in the nature and degree of impairment stems from both the nature of the acute insult and the subsequent development of neuroinflammation, diffuse axonal injury, diffuse vascular injury, and other secondary pathologies *(5–8)*.

Although cognitive rehabilitation can help patients develop strategies to adapt to their disability, such therapy has limited efficacy in remediating the memory deficits *(9)*. Given the profound unmet need facing patients with memory deficits related to acquired brain injury, we sought to examine whether closed-loop electrical stimulation can be effective in this patient group. As such therapies have yet to be validated, and because technology for chronic closed-loop stimulation is in its infancy, we sought to identify neurosurgical epilepsy patients with a significant prior history of TBI and test whether closed-loop neural stimulation could effectively boost memory in these patients.

Each year approximately 2,400 patients in the US undergo invasive electrocorticography monitoring for drug-resistant epilepsy, with the goal of localizing the seizure focus and planning a potentially-curative resective surgery. To localize seizures, neurosurgeons implant many electrodes that will prove to be outside the seizure onset zone; as such, these patients provide a unique window into the electrophysiology of memory and cognition. Additionally, neurologists often use electrical stimulation in such cases to map regions of eloquent cortex, so as to avoid resecting tissue vital to motor, language and memory function. During such cases researchers have also used electrical stimulation as a manipulative tool to study memory and cognitive processes. Investigation of open-loop stimulation protocols have at times demonstrated impaired memory *(10–12)* but in some cases, with careful targeting of specific tracts, these studies have also shown improved memory *(13–15)*.

Here we explored the possibility of using biomarker-guided, closed-loop electrical stimulation, to improve memory. Building upon a recent demonstration that closed-loop stimulation of lateral temporal cortex boosts memory for stimulated items we set out to evaluate whether this strategy would also work in a “therapy-based” setting, where stimulation would need to improve memory function throughout periods of potentially active stimulation. We further sought to validate this therapy in patients with a history of moderate-to-severe TBI. Among all patients undergoing invasive monitoring for resective surgery at six major epilepsy centers, we identified eight patients who met our TBI criteria (see *Materials and Methods*). Under an IRB approved protocol, and with an independent medical monitor reviewing patient safety data, we recruited these patients for a multi-session experiment involving memory testing, neural recordings, and closed-loop brain stimulation.

## Results

Consistent with prior work, our participants exhibited a moderate degree of memory impairment as determined by neuropsychological evaluation. Evaluations of memory included the Wechsler Memory Scale (WMS-IV), which was administered to all participants, and either the California Verbal Learning Test (CVLT-II) or the Rey Auditory Verbal Learning Test (RAVLT). Using the available indices of delayed recall, we constructed a composite measure that included the WMS-IV logical memory scale and the word-list recall measures from the CVLT-II and RAVLT (see *Methods*). Our participant group exhibited impaired memory, as seen in their average composite *z*-score of −1.01 (SEM = 0.31). This result aligns with prior TBI studies reporting a memory deficit of *z = -*0.82 on a similar index *(16)*.

During preliminary brain recording sessions we identified personalized biomarkers of successful memory encoding (spectral power at eight log-spaced frequencies between 6 and 180 Hz, see *Methods*) that we would use in later sessions to control closed-loop brain stimulation. To do this, participants first took part in computer-controlled memory tasks that would serve as “training” sessions for classifiers to learn personalized biomarkers indicative of successful or unsuccessful memory. Participants repeatedly studied lists of serially-presented nouns which they attempted to freely recall following a brief distractor task, designed to prevent active rehearsal (Fig. 1A, see *Methods* for details). This memory task mimics typical neuropsychological assessments (e.g., the CVLT mentioned above) but allowed us to relate multivariate neural activity captured by the 100+ clinical recording electrodes to patients’ behavioral performance.

**Fig. 1:**
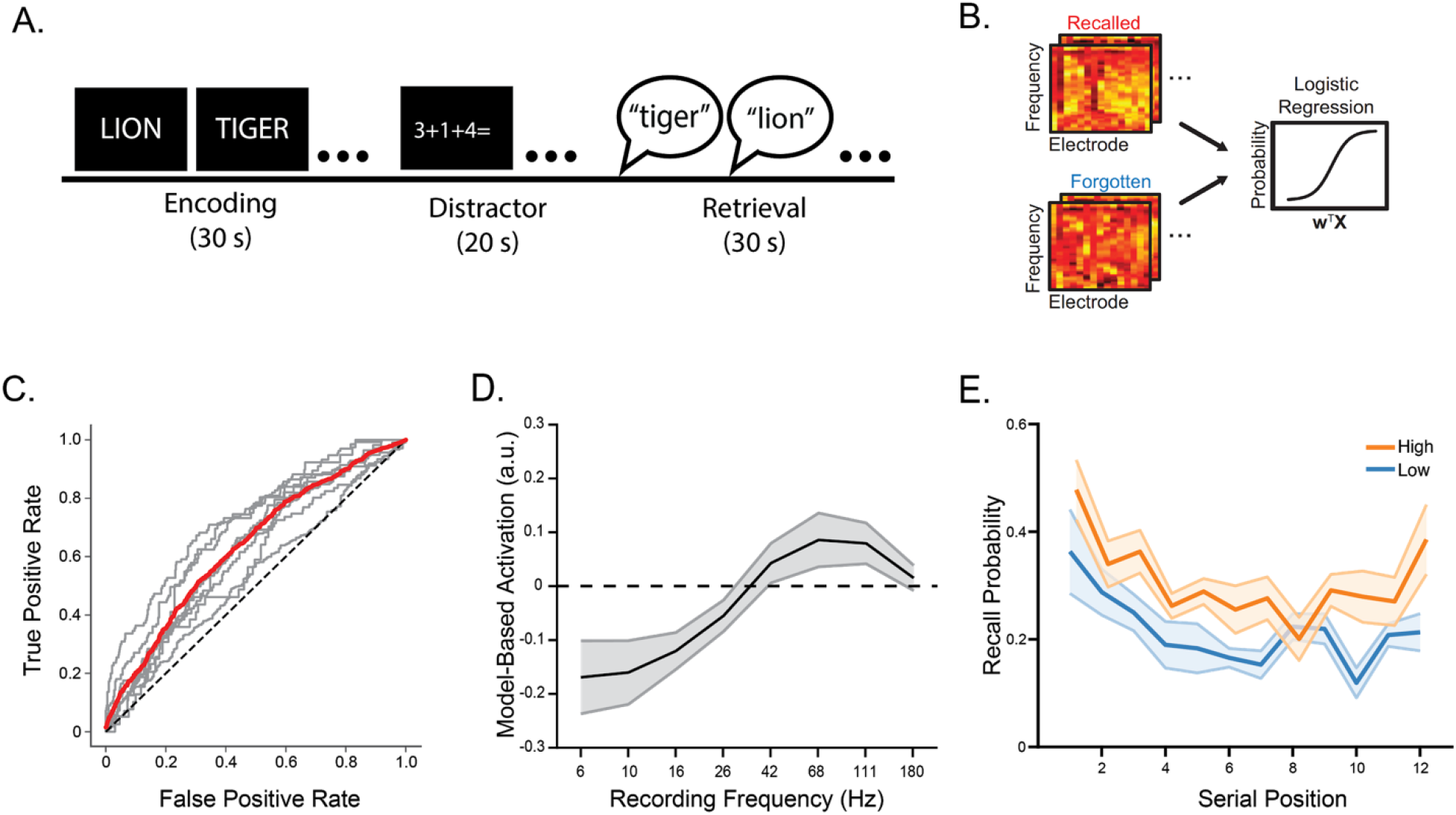
Assessing memory and decoding mnemonic success. (**A**) For each list, participants first studied a list of twelve sequentially-presented nouns, then performed an arithmetic-distractor task, and finally attempted to freely recall the studied items. Participants contributed data on 40+ study-test lists across 3+ experimental sessions. (**B**) We trained a logistic regression classifier to predict which items would be recalled on the basis of the neural activity (spectral power) measured at each electrode during memory encoding (vector labeled **x**). Training on hold-out sessions allowed us to estimate a weight matrix (**w**) associating brain activity with memory performance. (**C**) We examined the relation between the participant-specific neural classifier’s false-positive and true-positive rates, resulting in the ROC curves displayed here (mean ROC curve shown in red; overall classification performance well exceeding chance levels (AUC = 0.64; t_(7)_ = 6.40, p < 10^−3^). (**D**) Although we fit classifiers separately to each participant, the overall pattern derived by a forward model *(19)* revealed that increased high frequency and decreased low frequency activity marked periods of successful mnemonic processing. (**E**) Recall as a function of list position for items classified as in the top half (red) vs. bottom half (blue) of predicted recall, based on neural biomarkers. This shows the magnitude of the difference in predicted recall based on neural signals and demonstrates that the effect appeared consistently across list positions.

We constructed logistic-regression classifiers trained on spectral power extracted from each participant’s intracranial electroencephalography (iEEG) traces during word encoding, spanning frequencies from 6-180 Hz (i.e. the feature set). Using a “leave-one-session-out” cross-validation paradigm, classifiers performance was assessed only on held-out sessions of experimental data that were never seen by the training algorithm. This process yielded, for each subject, a weighted indication of how well each spectral feature could predict later memory. Previous studies have documented the success of mnemonic classification in much larger samples of >100 patients *(17, 18)*; here we show that this approach generalized to our TBI cohort (Fig. 1C). Periods of high classifier output – indicating a high probability of successful recall – were associated with increased power at high frequencies and diminished power at low frequencies (Fig. 1D). This effect appeared consistently across item positions within a list (Fig. 1E). These analyses show that classifier-based approach to predicting momentary changes in mnemonic function generalized well to our TBI cohort.

After building logistic-regression classifiers to decode variability in mnemonic success during record-only sessions, and meeting criteria for safe brain stimulation (see *Materials and Methods*), we advanced to the closed-loop stimulation experiment. Here, we again administered a series of delayed-recall lists, randomly assigning each list to either a stimulation or non-stimulation condition. Within stimulation lists, our algorithm decoded mnemonic success in real time using the previously constructed classifiers and triggered 500 ms bouts of 200 Hz, 0.5 mA stimulation when the classifier signaled that a patient’s memory performance dipped below their predicted average (Fig. 2A). We restricted stimulation to contacts on the left lateral temporal cortex, based on evidence from a prior study *(20)*. Additionally, a recent analysis of stimulation frequencies *(21)* indicated that 200 Hz stimulation more strongly modulated high-frequency activity -- an established biomarker of cognition function in human hippocampus and neocortex *(22–24)*.

**Fig. 2:**
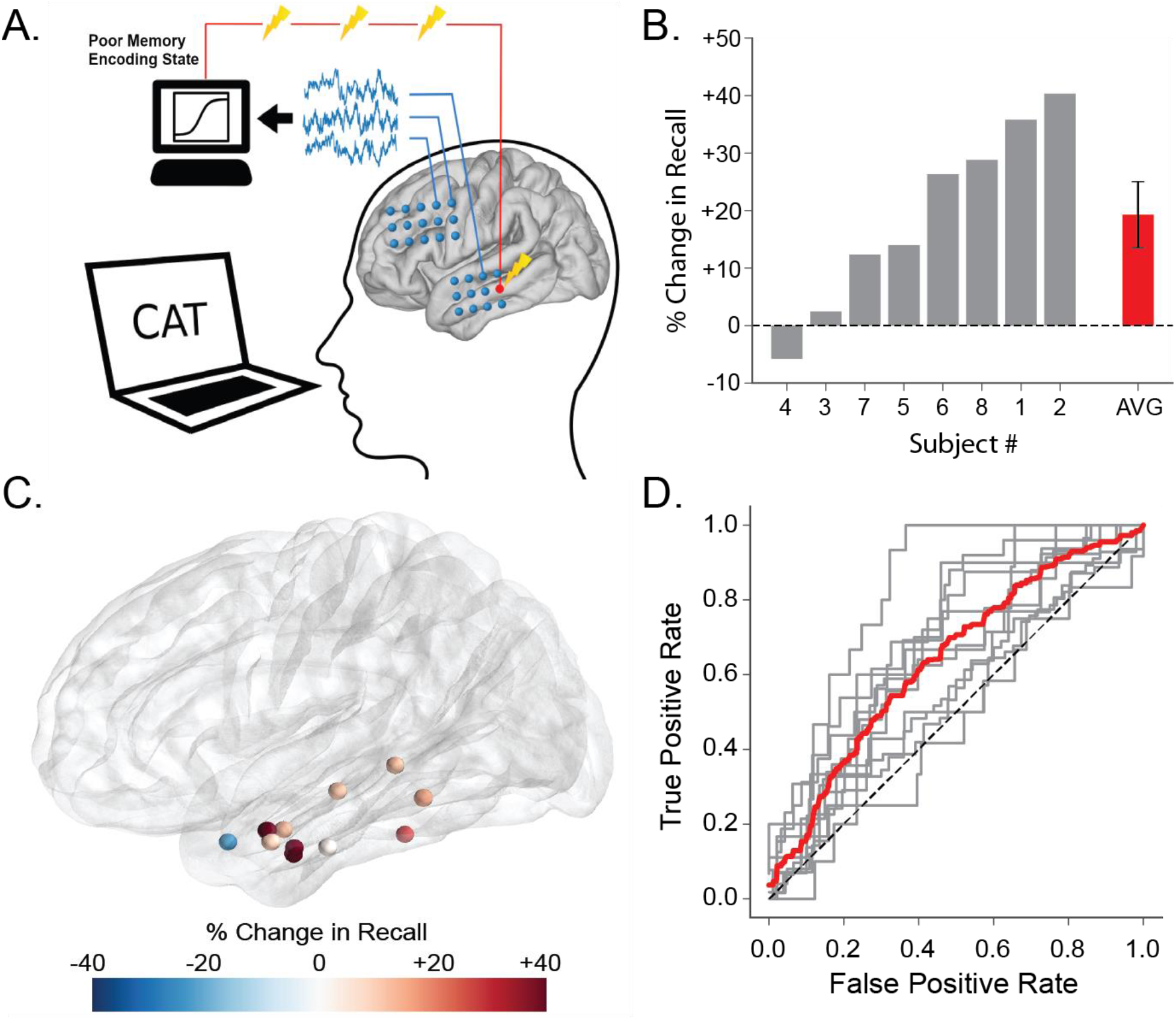
Closed-loop stimulation and memory performance. (**A**) While the patient views a word during the memory encoding phase of the delayed recall task (Fig 1A), we applied the logistic-regression classifiers trained in earlier record-only sessions (Fig.1B) to predict mnemonic success. Our algorithm triggered 500-ms bouts of 200 Hz stimulation to the lateral temporal cortex whenever the estimated probability of recall dropped below 0.5 (**B**) Participant-level memory improvement/impairment. Participants experienced an average 19% increase in recall performance (red) with seven of eight patients experiencing some positive effect of closed loop stimulation (t_(7)_ = 2.92, p = 0.012). (**C**) Stimulation targets for each unique stimulation site (n=11 sessions) rendered on an average brain surface; sphere shading indicates the percent change in word recall performance. Targets were distributed across the superior, middle, and inferior temporal gyrus, though largely located in the middle temporal gyrus. (**D**) Classifier performance. Receiver operating characteristic (ROC) curves showing performance of a record-only classifier tested on No-Stim lists during the closed loop sessions (gray lines, n=11 sessions; red line, average ROC). The overall AUC across sessions reliably exceeded chance levels (t_(10)_ = 4.82, p < 10^−3^; see *Methods*).

Our primary question was whether a fully closed-loop therapy could improve memory for entire lists during which only some items are stimulated; prior studies considered stimulation’s effect on individual items, leaving open the possibility that helping some items came at the expense of hurting others. We thus examined recall rates in lists with closed-loop stimulation and compared those to lists in which stimulation was disabled (non-stimulation, or “sham” lists). Participants recalled reliably more items on the stimulation than on the sham lists (25.2% vs. 21.1%; two-tailed paired t-test, *t*_*(7)*_ *= 3*.*36, p = 0*.*012*). Fig. 2B illustrates the degree of memory improvement (or impairment) for each of our eight participants. Here we quantified stimulation-related memory improvement (or impairment) as the mean difference in stimulation and non-stimulation list recall rates (determined for each session), divided by the average non-stimulation recall rate across participants. On average, closed-loop LTC stimulation improved recall by 19% (Fig. 2B). Our algorithm triggered stimulation an average of 6.5 times during each stimulation list.

We applied a hierarchical linear mixed-effects model to account for the effect of multiple unique stimulation targets (three participants took part in an additional stimulation session with a different target location yielding a total of 11 unique targets, Fig. 2C), as well as the effect of list position within each session (to control for variability in the randomly assigned positions of stimulation and non-stimulation lists). This model relates recall performance on each list to the presence of stimulation during each list and the list position within each session. Recall performance often varies over the course of a session *(25)* leading to a potential bias if the random assignment of stimulation conditions to list positions results in one condition occurring in more favorable list positions within a session. A positive relation (β parameter) between recall rate and stimulation indicates higher memory performance during lists where poor-biomarker states triggered stimulation (as compared with the non-stimulation, “sham” lists). We observed a main effect of closed-loop stimulation (*β = 0*.*043;* likelihood-ratio test, *χ*^*2*^_*(1)*_ *= 5*.*5; p = 0*.*019*) as well as an effect of list position within the session (*β = −0*.*004;* likelihood-ratio test, *χ*^*2*^_*(1)*_ *= 9*.*7; p = 0*.*0018*). These results indicate that the delivery of closed-loop LTC stimulation improved list-level recall even when controlling for list position and statistically modeling the effect of repeated sessions.

Seven of the eight participants receiving closed-loop stimulation experienced improved memory on stimulation lists, but the degree of improvement varied considerably across participants (Fig. 2B). Although our sample size did not permit a quantitative analysis of individual differences in response to therapy, two variables deserve mention. Stimulation targets for the eleven sessions lay across variable regions of the superior, middle, and inferior temporal gyrus, due to clinical constraints. Fig. 2C illustrates each stimulated target on an average brain along with its modulatory effect on recall, indicated by the shading of each sphere. Classifier generalization also varied across stimulation sessions (Fig. 2D) which would increase variability in the effectiveness of any closed-loop algorithm. Elucidation of these and other potential mediating factors will require larger studies and explicit manipulations.

## Discussion

Direct electrical brain stimulation has emerged as a viable therapeutic tool to rescue function in patients with progressive neurological disorders, most prominently refractory Parkinson’s disease and essential tremor *(26, 27)*, and more recently, intractable epilepsy *(28, 29)*, depression *(30)*, and obsessive-compulsive disorder *(31)*. The lack of effective pharmaceutical therapies for many neurological and psychiatric conditions has prompted researchers to investigate the potential utility of electrical stimulation in treating a host of other indications, including depression, eating disorders, and addiction *(32)*.

The present study offers a proof-of-concept for a closed-loop brain stimulation strategy to treat memory loss in TBI. By first training logistic regression classifiers on spectral power features observed during memory encoding, we demonstrated that these classifiers could accurately predict subsequent memory even before a subject engaged in overt recall. We further showed that these same classifiers could form the basis of a closed-loop stimulation algorithm, in which electrical stimulation to the temporal cortex was delivered during predicted memory lapses. Recapitualting earlier work in a heterogenous epilepsy cohort *(20)*, this approach yielded an average 19% improvement in recalled items across our TBI group, with individual positive effects observed in seven of the eight patients.

To gauge the meaningfulness of a 19% memory boost, we need to consider the degree of impairment in this cohort. Among patients with moderate-to-severe TBI, one typically finds that delayed recall performance is 0.82 standard deviations below the average performance of age-matched controls *(16)*. Based on the distribution of performance of TBI-afflicted individuals in delayed-free recall, the 19% improvement observed in our study implies a 0.44 standard deviation increase. Our results therefore suggest a theoretical reduction in the burden of this type of memory impairment by 53.6% in these patients. By demonstrating therapeutic efficacy in patients who have both a history of moderate-to-severe TBI and documented memory impairment we hope our findings will accelerate the development of technologies for patients with acquired brain injuries, which could restore some degree of their lost memory function as they attempt to rebuild their post-injury lives.

While these findings are encouraging, more work remains before this technique can be applied in a therapeutic setting. First, the electrophysiological responses – not just behavioral responses – to brain stimulation must be explored to better understand the neural mechanisms underlying improved memory performance. Several studies have recently characterized spectral responses to direct brain stimulation, with these early results suggesting that both low and high-frequency oscillations can be induced by exogenous pulse trains, depending partly on the local tissue architecture of the stimulation site *(33)*. Relatedly, it remains an open question as to how the underlying connectivity profile of a stimulation site interacts with consequent change in behavior – the structural and functional connectivity of a stimulation site with the medial temporal lobe, and other regions of the core memory network, is a point of particular interest and should be characterized in future work *(34)*.

The long-term efficacy of chronically indwelling electrodes for memory enhancement is entirely uncharacterized, though addressing this question will be critical to the eventual deployment of brain stimulation as a memory therapeutic in TBI patients. The use of direct electrical stimulation in Parkinson’s and epilepsy, among other neuropsychiatric conditions, suggests that chronic implantation is safe and effective *(35)*. However, chronic modulation of the neural circuitry underlying episodic memory could pose additional challenges, including adaptive changes in neural responses to stimulation.

Finally, our study constitutes a rare case in which direct brain recordings and stimulation can be ethically carried out in a group of TBI patients. As such, our study population is small, and likely does not capture the full heterogeneity of TBI pathologies. Through continued multi-institutional efforts, we hope that future work can extend these results to larger samples, and thus more precisely determine the efficacy of therapeutic brain stimulation across a range of underlying traumatic neuropathologies.

## Materials and Methods

### Study Design

Based on prior published work in an independent data set *(20)*, we hypothesized that closed-loop stimulation of lateral temporal cortex in human subjects with a history of memory dysfunction and traumatic brain injury would provide a boost in recall on an episodic memory task during trials receiving stimulation as compared to trials without stimulation.

### Research subjects

We recruited eight patients (7 male, 1 female; mean age 44.5 +/- 11 SD) with intractable epilepsy who were undergoing seizure monitoring and localization using implanted intracranial electrodes. We determined the sample size in advance and did not perform group analyses until we completed data collection. We identified patients as having a history of moderate-to-severe TBI based on criteria established by expert neurologists at the University of Pennsylvania (Dr. R. D-A) and University of Texas Southwestern (Dr. K. D.), as follows: a reported history of significant head injury accompanied by either prolonged loss of consciousness (>30 minutes), post-traumatic amnesia, or imaging results compatible with moderate-to-severe traumatic brain injury. We confirmed left-language dominance based on functional magnetic resonance imaging (fMRI) and/or WADA testing in six of these patients; the remaining two patients did not have specific data confirming language dominance, but one was right-handed and other self-reported as being ambidextrous. The enrolled patients participated at the following collaborating hospitals: Dartmouth-Hitchcock Medical Center (Hanover, NH), Emory University Hospital (Atlanta, Georgia), and University of Texas Southwestern Medical Center (Dallas, TX), with the University of Pennsylvania (Philadelphia, PA) serving as the Data Coordinating Center. This research was part of a multi-center project designed to assess the effects of electrical stimulation on memory-related brain function. Institutional review boards approved the study protocol at the respective institutions, and each participant gave written informed consent after the nature and possible consequences of the study were explained.

### Experimental Design

All enrolled participants underwent the same experimental protocol: multiple sessions of a non-stimulation behavioral task followed by one or more sessions of a closed-loop stimulation task (see details in following subsections). Each participant served as their own control within each stimulation task session (control trials receiving no stimulation). Patients were aware that stimulation could be delivered throughout the closed-loop stimulation task but were blinded to the specific trials containing stimulation. We included all closed-loop stimulation task sessions performed by the participants in the reported results and did not perform group analyses until all data were collected.

### Behavioral task

Each participant performed a delayed verbal free recall task (Fig. 1A) on a laptop, in which they studied lists of displayed items for later verbal recall. Each list comprised 12 words selected without replacement from a pool of nouns; the word pool consisted of items belonging to 25 semantic categories (e.g. beverages, kitchen appliances, zoo animals). Each list of 12 items consisted of four unique words drawn at random (and without replacement) from each of three randomly-selected categories. Participants contributed up to 25 study-test trials per session, plus a practice trial discarded in subsequent analyses. Each trial consisted of three main phases: encoding, distractor, and recall (Fig. 1A). Following a ten-second countdown period, each trial began with the encoding phase, in which the computer displays each item individually for 1600 ms followed by a randomly jittered 750-1000 ms blank inter-stimulus interval (ISI). After viewing the final item of the list, participants entered a distractor phase (20 seconds), in which they typed responses to a series of simple arithmetic problems, receiving correct/incorrect feedback through an audio tone. Following the distractor phase, a brief auditory tone cued participants to speak aloud as many items as possible from the most recent viewed list (30 seconds), in any order, with vocal responses digitally recorded and later manually annotated for analysis. Participants performed the categorized delayed free recall task without brain stimulation (i.e. “record-only”) or with closed-loop brain stimulation (see details below). Two of the eight participants performed record-only sessions of a nearly-identical uncategorized variant of the delayed free recall task in addition to the categorized variant described above, in which the word pool consisted of high frequency nouns. In prior work, analyses of behavioral and electrophysiological data across these two task variants have revealed nearly identical biomarkers relating to successful memory encoding and retrieval *(36)*.

### Data collection and processing

The neurosurgical team implanted minimally-invasive stereoelectroencephalography (sEEG) depth electrodes (AdTech Medical Instrument Corporation, PMT Corporation, DIXI Medical) within the brain parenchyma to collect electrophysiological data (EEG) to best localize epileptogenic regions, with specific placement of each electrode planned to support each patient’s individual clinical care. The External Neural Stimulator (ENS) (Medtronic, Inc.) recorded EEG signals (microvolts; sample rate = 2000 Hz) using a bipolar reference scheme, consisting of pairs of immediately adjacent contacts on the implanted sEEG electrodes. During testing sessions, the laptop recorded behavioral responses (vocalizations, key presses), synchronized to the ENS-recorded EEG via transmitted network packets. In a first pre-processing step, a 5 Hz band-stop fourth order Butterworth filter (centered on 60 Hz) removed line noise from the recorded EEG signals. Then, we performed a spectral power decomposition on the time-series data at 8 frequencies from 6-180 Hz, logarithmically-spaced, using Morlet wavelets (wave number = 5) for time windows from 0 to 1366 ms relative to word onset, including a mirrored buffer (length = 1365 ms) before and after the interval of interest, in order to avoid convolution edge effects. Finally, we log-transformed the resulting time-frequency data, averaged over the time interval, and *z*-scored within session and frequency across item presentations.

### Localization of electrodes to anatomy

We performed a patient-specific parcellation of cortical surface regions according to the Desikan-Killiany atlas using Freesurfer *(37)* on pre-surgical volumetric T1-weighted magnetic resonance imaging (MRI) scans. We performed an additional volumetric segmentation of the whole brain cortical surface and medial temporal lobe was performed on the T1-weighted scan and a high-resolution hippocampal coronal T2-weighted scan using Advanced Normalization Tools (ANTS) *(38)* and Automatic Segmentation of Hippocampal Subfields (ASHS) multi-atlas segmentation methods *(39)*. We derived coordinates of the radiodense electrode contacts from a post-implant CT using custom software (Voxtool, https://github.com/pennmem/voxTool), and co-registered with the T1 and T2 MRI scans using ANTS.

### Classification of mnemonic success

Each participant took part in multiple sessions of a delayed free recall memory task (Fig. 1A). The record-only sessions provided behavioral data and EEG time-series data upon which to train a participant-specific multivariate logistic regression classifier to identify patterns of brain-wide neural activity during memory encoding that predicted recall success *(17, 18, 22, 36)*. The classifier utilized recorded EEG spectral power as features for training, measured during successful and unsuccessful memory encoding and memory retrieval event epochs from prior record-only sessions (Fig. 1B). Encoding event epochs spanned the time 0-1366 ms relative to item presentation, and retrieval event epochs spanned −525-0 ms relative to either a valid recall or an unsuccessful search period. Valid recalls did not include repeated items and intrusions of non-list items. We defined a deliberation period as a 525 ms interval in which no prior vocalizations occurred within 1000 ms from onset and no subsequent recall within 2000 ms. We also excluded time periods within the first and last 1000 ms of the entire retrieval interval. For each valid pre-retrieval epoch, we identified a “matched” unsuccessful search deliberation epoch on another list, selecting the epoch from the closest list within the session whose timing within the recall period matched that of the valid recall and no vocalization took place. Given the potential imbalance between encoding and retrieval events, we inversely weighted the penalty parameter according to the class imbalance *(40)*, and computed class weights for each class as

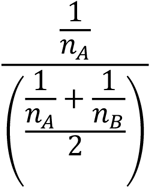

where *n*_*A*_ is the number of events class *A* events, and *n*_*B*_ is the number of class *B* events. Encoding/retrieval class observations had a fixed value weighting of 2.5 *(17)*.

For each participant, we measured classifier performance on the training dataset using a leave-one-session-out (LOSO) cross-validation procedure, by measuring the area under the curve (AUC) of the receiver operating characteristic (ROC) curve. The AUC of the ROC relates the ratio of the true positive rate (correctly classified as later recalled) and false positive rate (incorrectly classified as later recalled) across classification thresholds (Fig. 1C). A two-tailed, one-sample *t*-test versus chance level AUC of 0.5 (alpha = 0.05) tested for significance of trained classifier AUCs across participants. To measure generalization performance of a participant’s classifier to the closed-loop session, we used the same AUC metric calculated using the No-Stim lists as the evaluation dataset (Fig. 2D), testing for significance using the above described t-test across closed-loop stimulation sessions. To assess the relative importance of different frequency features to the classifier’s performance (Fig. 1D), we calculated a forward model for each participant based on the data covariance matrix and the weights of fitted classifier. We also used the classifier’s outputs on the training data to separate items into low and high probability of memory success based on the median of the distribution of classifier outputs across all items (Fig. 1E). This allowed us to characterize how participants’ actual memory performance compared with the predictions of the model.

### Closed-loop stimulation task and procedure

In subsequent closed-loop stimulation sessions, the closed-loop system used the multivariate logistic regression classifier to decode the probability of recall from neural activity on-line during the encoding phase of the task (Fig. 2A). Subjects first performed one No-Stim practice list, followed by 25 additional Stim and No-Stim lists. Lists 1-3 were used as a No-Stim baseline for normalizing the classifier features; lists 4-25 consisted of 16 lists of Stim and 6 lists of No-Stim conditions, randomly distributed. Concurrent with the free recall task, the closed-loop system calculated spectral power features on recorded data from 0-1366 ms relative to item presentation: during Stim lists, if the predicted probability of recall was below 0.5 (i.e. a poor memory encoding state), the system immediately triggered stimulation. On No-Stim lists, stimulation was not delivered. During each stimulation event, electrical current passed through a single pair of adjacent electrode contacts, as charge-balanced biphasic rectangular pulses (pulse width = 300 μs) at 200 Hz frequency, for 500 ms. Each stimulation session began with the determination of a safe stimulation amplitude, in which experimenters initially triggered stimulation trains at the above parameters at a low, floor amplitude chosen by the monitoring clinician, who observed the patient’s EEG for stimulation-induced afterdischarges. With the clinician’s approval, the amplitude of additional stimulation incrementally increased, until reaching either the target amplitude or a safe maximum amplitude. Target amplitudes were below the afterdischarge threshold and below accepted safety limits for charge density; in this study, all participants received stimulation at 0.5 milliamperes. As the neurosurgical team determined electrode implantation sites on a case-by-case basis to address each patient’s specific care, we used a combination of anatomical and functional information to select stimulation sites. We prioritized electrodes in lateral temporal cortex (LTC), in particular the middle portion of the middle temporal gyrus, where previously published results have indicated a stronger beneficial effect *(20)*.

### Statistical analysis of the effect of stimulation on memory

To assess the effect of stimulation on memory performance, we compared participant recall rates on Stim lists and No-Stim lists. As standard practice, we excluded the practice list and the first three baseline (non-stimulation) lists from these analyses. For each session, we calculated the mean percentage of recalled words for the Stim and No-Stim list conditions (i.e. number words recalled divided by total number of words). To account for inter-session baseline differences in recall, we calculated the difference in individual session Stim and No-Stim recall rates, normalized by the mean No-Stim recall rate across all sessions, and tested for significance using a two-tailed paired t-test (alpha = 0.05).

We further estimated the effect of stimulation on memory performance using a hierarchical linear mixed effects model that took account of the varying numbers of closed-loop sessions across participants and the effect of list position within each session. Here, we analyzed recall percentage in the model, with Stim/No-Stim and list number (position with the session) as fixed effects and session nested in participant as a random intercept effect. A likelihood-ratio chi-squared test (alpha = 0.05) evaluated the significance of the fixed effects by comparing the performance of the full model to that of a reduced model without the fixed effect in question.

## Data Availability

We have posted all de-identified data, including EEG, localization information, and behavioral data, to the cognitive electrophysiology portal at memory.psych.upenn.edu. Correspondence and requests for materials should be addressed to M.J.K.

http://memory.psych.upenn.edu/

## Acknowledgments

The authors express their gratitude to the patients who selflessly volunteered to participate in these studies. They acknowledge Linley Robinson, Robert Quon, and Lou Blanpain for their role in data collection. They acknowledge Dr. Daniel Rizzuto for contributions to experimental design and the Medtronic team, led by Dr. Tim Denison, for their contributions to the development of the ENS neural recording and stimulation platform.

## Author contributions

M.J.K, P.A.W., and Y.E., designed experiments. M.J.K, P.A.W. and Y.E. analyzed the data. P.A.W., R.A-Z. performed and coordinated data collection. B.C.L., B.C.J., R.E.G. recruited participants and provided general assistance. K.D. and R.D-A. provided expert guidance regarding traumatic brain injury history and criterion. M.J.K., P.A.W, and R.D.A. wrote the manuscript. All authors provided feedback on the manuscript.

